# Poorly known aspects of flattening the curve of COVID-19

**DOI:** 10.1101/2020.06.09.20126128

**Authors:** Alain Debecker, Theodore Modis

## Abstract

This work concerns the too-often mentioned flattening of the curve of COVID-19. The diffusion of the virus is analyzed with logistic-curve fits on the 25 countries most affected at the time of the writing and in which the diffusion curve was more than 95% completed. A negative correlation observed between the final number of infections and the slope of the logistic curve corroborates a result obtained long time ago via an extensive simulation study. There is both theoretical arguments and experimental evidence for the existence of such correlations. The flattening of the curve results in a retardation of the curve’s midpoint, which entails an increase in the final number of infections. It is possible that more lives are lost at the end by this process. Our analysis also permits evaluation of the various governments’ interventions in terms of rapidity of response, efficiency of the actions taken (the amount of flattening achieved), and the number of days by which the curve was delayed. Not surprisingly, early decisive response proves to be the optimum strategy among the countries studied.

## 1. Introduction

As early as in 1925 Alfred J. Lotka demonstrated that manmade products diffuse in society along S-shaped patterns similar to those of the populations of biological organisms.[1] Since then S-curve logistic descriptions have made their appearance in a wide range of applications from biology, epidemiology, and ecology to industry, competitive substitutions, art, personal achievement and others.[2-5] The most fascinating aspect of S-curve fitting is the ability to predict from early measurements the final ceiling. This very fact, however, constitutes also the fundamental weakness and the major criticism of these predictions because logistic fits on early data can often accommodate very different values for the final ceiling. Obviously, the more precise the data and the more of the S-curve range they cover, the more accurate the determination of the final level but, unfortunately, at the same time, the less interesting this prediction becomes.

In the mid-1980s we began studying the sales of computers with S-curves.[6] It soon became obvious that it was of crucial importance to quantify the uncertainties involved in the determination of the parameters of our logistic fits. As a consequence we carried out an extensive simulation study aiming to quantify the uncertainties involved in fitting data with logistic curves, which was published in 1994.[7] The study was based on some 35,000 S-curve fits on simulated data, smeared by random noise and covering a variety of conditions. The fits were carried out via a *χ*^2^ minimization technique. The study produced look-up tables and graphs for determining the uncertainties expected on the three parameters *M, α*, and *t*_0_of the logistic function:

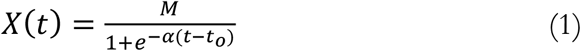

In addition, our study established correlations between these three parameters. In particular, a negative correlation was found between the level of the final ceiling *M* (the niche capacity) and the rate of growth (the slope *α*.) This observation is another manifestation of a correlation—or trade-off—often encountered in a various disciplines and situations. For example, in chemical reactions there exists such a trade-off between the reaction rate (the time to complete a reaction) and the yield of the reaction. A typical case in point is the synthesis of ammonia (the Haber Bosch process.) Another example has been pointed out more recently between the power (i.e. charging/discharging rate) and the capacity in batteries.[8] A classical case of this kind of trade-off is between power and efficiency in heat engines. It has been described in an article by Naoto Shiraishi, Keiji Saito, and Hal Tasaki, where they explain that the origin of such trade-offs is the existence of thermodynamic constraints.[9]

It is not surprising to find this correlation in biological systems because they too constitute some kind of heat/chemical engines. In fact the growth rate vs yield trade-off has long been known in biological systems. The growth rate versus niche capacity, and the efficiency versus power (or productivity) trade-offs for biological systems have been treated by Alfred J. Lotka in an article published in 1922 with title “Contribution to the Energetics of Evolution.” He wrote:

> “Where the supply of available energy is limited, the advantage will go to that organism which is most efficient, most economical, in applying to preservative uses such energy as it captures. Where the energy supply is capable of expansion, efficiency or economy, though still an advantage, is only one way of meeting the situation, and, so long as there remains an unutilized margin of available energy, sooner or later the battle, presumably, will be between two groups or species equally efficient, equally economical, but the one more apt than the other in tapping previously unutilized sources of available energy.”[10]

The onslaught of the COVID-19 in early 2020 triggered widespread interest in the use of S-curves to describe the diffusion of the virus in different countries. At the same time, the concept of flattening the bell-shaped curve of the rate of diffusion acquired importance and urgency. From the beginning of the COVID-19 epidemic governments around the world undertook efforts—varying in urgency and efficiency from one country to another—in order to flatten the bell-shaped curve of the rate of infections in their country. The idea behind this effort was to slow down the rate of the disease’s spreading thus permitting hospitals to handle the increasing numbers of patients in need of intensive care. The kind of measures governments took and the effectiveness of their application influenced the rate of growth *α* and distorted the bell-shaped curve into asymmetric distributions. The magnitude of this distortion reflects the kind of measures taken and their effectiveness. Typically, a rapidly imposed strict lockdown decreased the value of *α* abruptly causing an early inflection point on the S-curve. Other measures, like social distancing, influenced the value of *α* differently. As a consequence we saw a variety of distorted S-curves in countries around the world depending on the actions taken by their governments.

People at the time were not particularly concerned with how this flattening may impact the final number of infections. The implicit assumption all along was that this number would probably also decrease with the various measures taken. However, the negative correlation *M* between and *α* mentioned earlier suggests otherwise. With many countries now having completed the first or the main wave of the virus diffusion there it becomes possible to study experimentally the relationship between *M* and *α* in various countries around the world.

The work described here uses the parameters from S-curve descriptions of the COVID- 19 diffusion in the countries most affected at the time of the writing (mid-May 2020.) The conclusion is that the number of people finally infected by the virus was probably significantly increased as a consequence of the way some curves were flattened, which raises questions about the wisdom of those measures and the way they were taken.

## 2. The Number of Infections versus the Rate of Diffusion

The negative correlation between parameters *M* and *α* of Equation (1) established in our simulation study mentioned earlier is shown in Figure 1, which reproduces Fig. 7(a) from that article. In particular, we see that for a 10% drop in the value of, say from 1.1 to 1.0 we could expect an increase in the value of *M* as much as a factor of 2.

**Figure 1.**
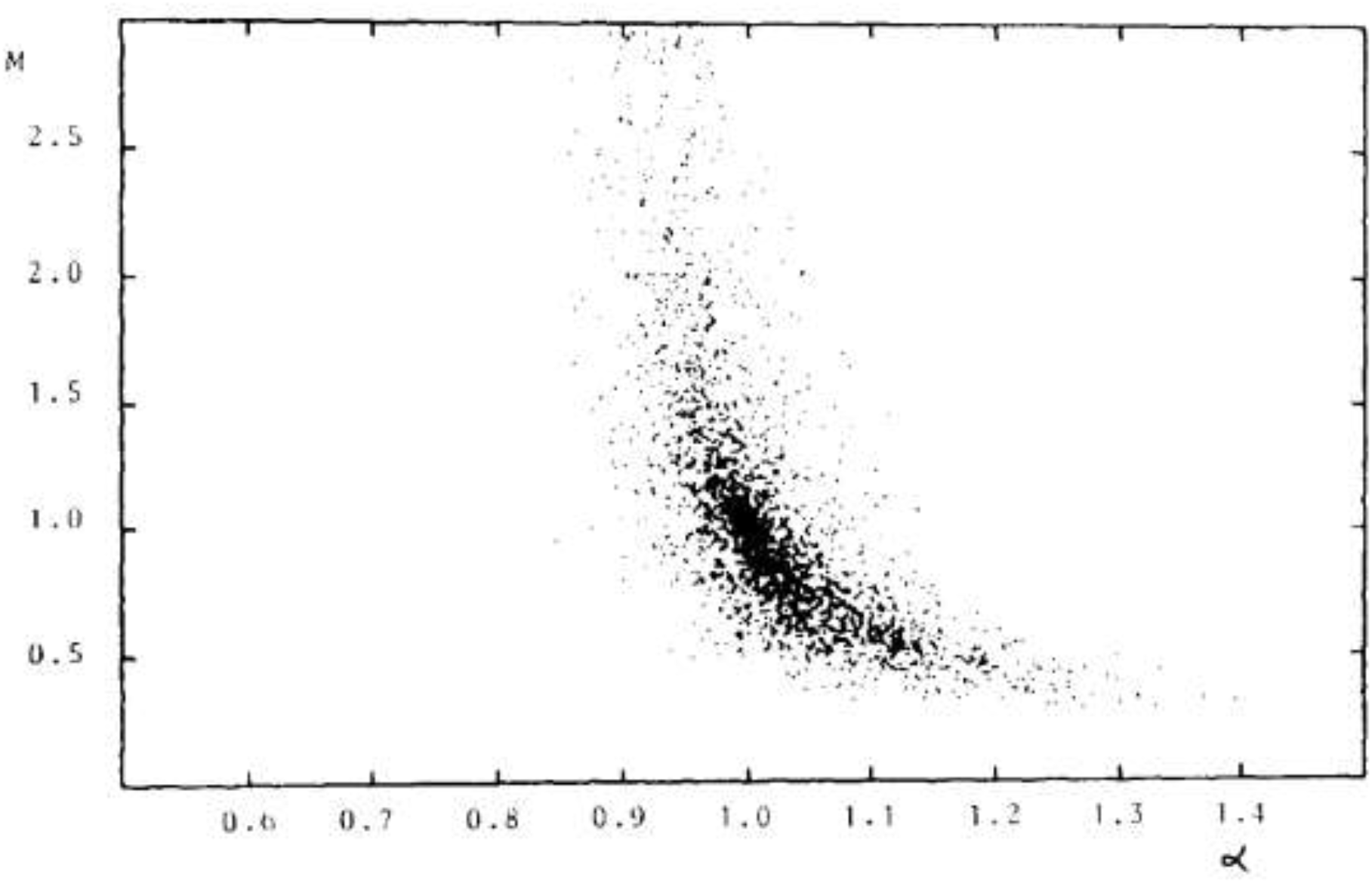
The correlation between *M* and *α* reproduced here from the publication of Debecker and Modis. Every dot represents a logistic fit on simulated data (with added noise) that cover the early 20% of a logistic S-curve.

With this correlation in mind we became motivated to search experimentally for possible correlation between the number of infections and the rate at which they diffused in different countries. We fitted logistic S-curves—namely Equation (1)—on the diffusion data of COVID-19. Among the most affected countries we selected those that had completed their S-curve (of the first or the main wave) to more than 95%. The twenty-five countries thus retained represented 70% of the world’s infections at the time of the writing (mid-May 2020.) The most affected country, USA, is only partially analyzed because its final curve was far from being completed at the time of the writing.

The data come from four different sources and they have been cross-verified for each country by at least two of these sources.[11-14] For China and the USA, the regions particularly affected, namely Hubei Province and New York respectively, have been separated from the rest of the country; so there are a two graphs for China but only one (New York) in the USA because the rest of the country there was nowhere near completion of its S-curve at the time of the writing. Fitted curves, parameters, and other data for all countries are shown in the Appendix.

In Figure 2 we see a scatter plot depicting *M* versus *α* for the S-curves of the twenty-six graphs in the Appendix; it corroborates a negative correlation qualitatively similar to the one shown in Figure 1.

**Figure 2.**
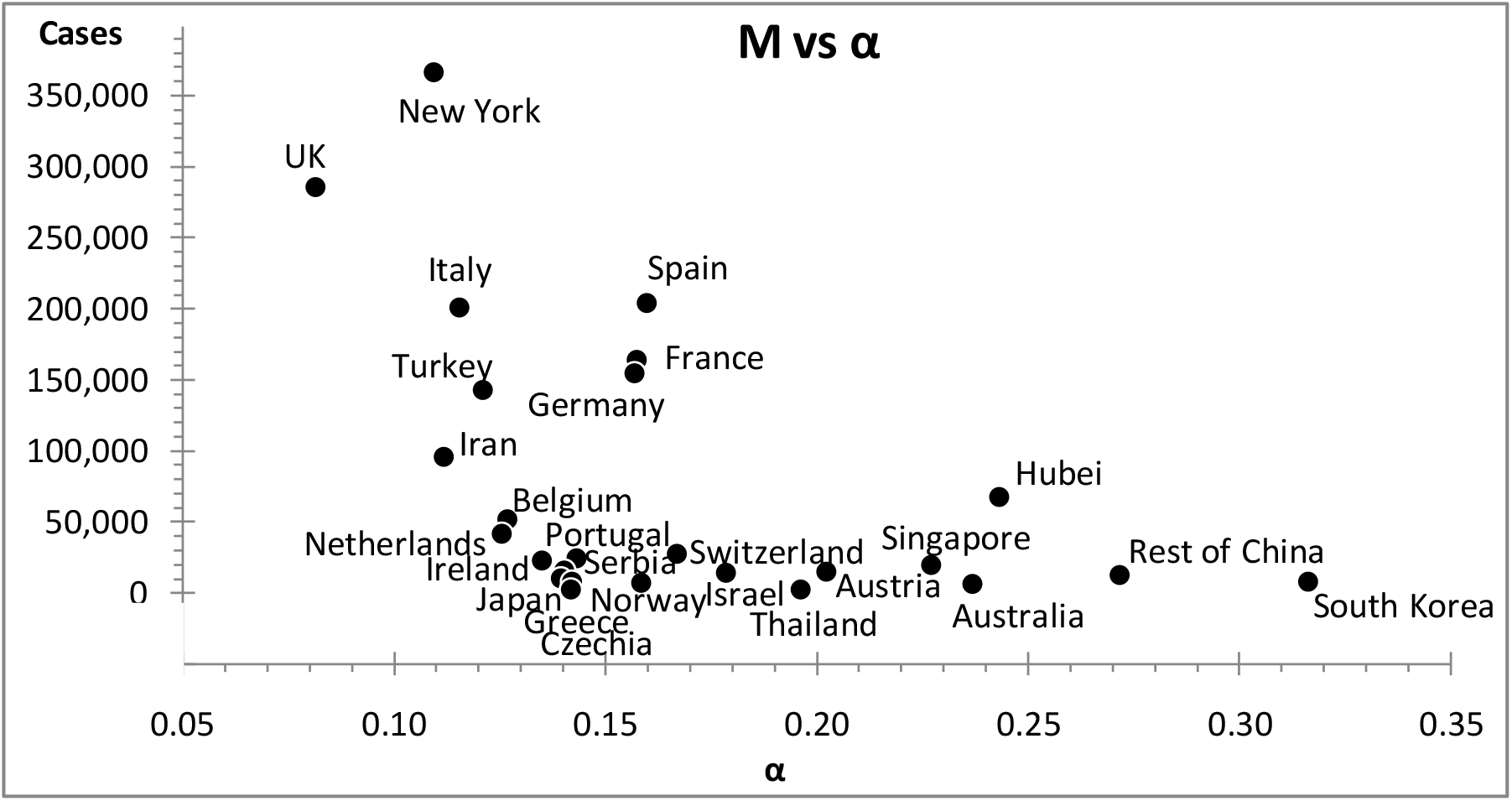
A scatter plot of *M* versus *α* for the most affected countries that completed their S-curves; as a rule the flatter the slope of the S-curve the greater the number of confirmed cases.

## 3. On the Actions Taken

In response to the pandemic different governments took different actions in two directions, to ameliorate the existing health system (building hospitals, distributing patients toward less affected regions, procuring respiratory ventilators, etc.) but also to reduce the diffusion rate (lockdown, social distancing, the use of masks, restriction on travel and social gathering, etc.) The second type of actions impacted directly the rate of diffusion, which kept changing differently in different countries depending on the rapidity and the effectiveness of the actions taken. As a consequence the logistic S-shaped pattern of the overall number of infections, and the corresponding bell-shaped pattern of the number of daily cases, got distorted to a greater or lesser extent. Rapid effective action, like the case of South Korea, results in a rather symmetric curve, whereas delayed/inefficient actions resulted in a distorted curve with the trailing side of the bell-shaped curve prolonged. More extreme such cases, like the rest of the USA (outside New York), Sweden, and Poland, saw an extended flattening of the bell-shaped curve, which instead of a peak displayed a plateau prolonged over months. We made no attempt to fit overall S-curves to these countries.

Despite the fact that the parameters we determine from the overall logistic fits carry negligible uncertainties—consequence of the fact that these S-curves are practically completed—their values are somewhat of a compromise because these parameters give the best but not always a textbooklike logistic description of the data. For example, the slope *α* is in fact an average value over several *α*’s, resulting from a sequence of different government actions as they went through imposing and later lifting lockdowns, social distancing, restrictions on social gatherings, and the like. For each country we distinguish *α*1as the slope of the very early logistic curve, before any government action took effect. We determined these early slopes separately by fitting the datapoints only up to the maximum daily rate—often coinciding with the so-called peak—which typically occurs around fifteen days following the establishment of a lockdown. In Figure 3 we see that three quarters of the countries we studied reached a peak within 8 to 19 days from lockdown start.

**Figure 3.**
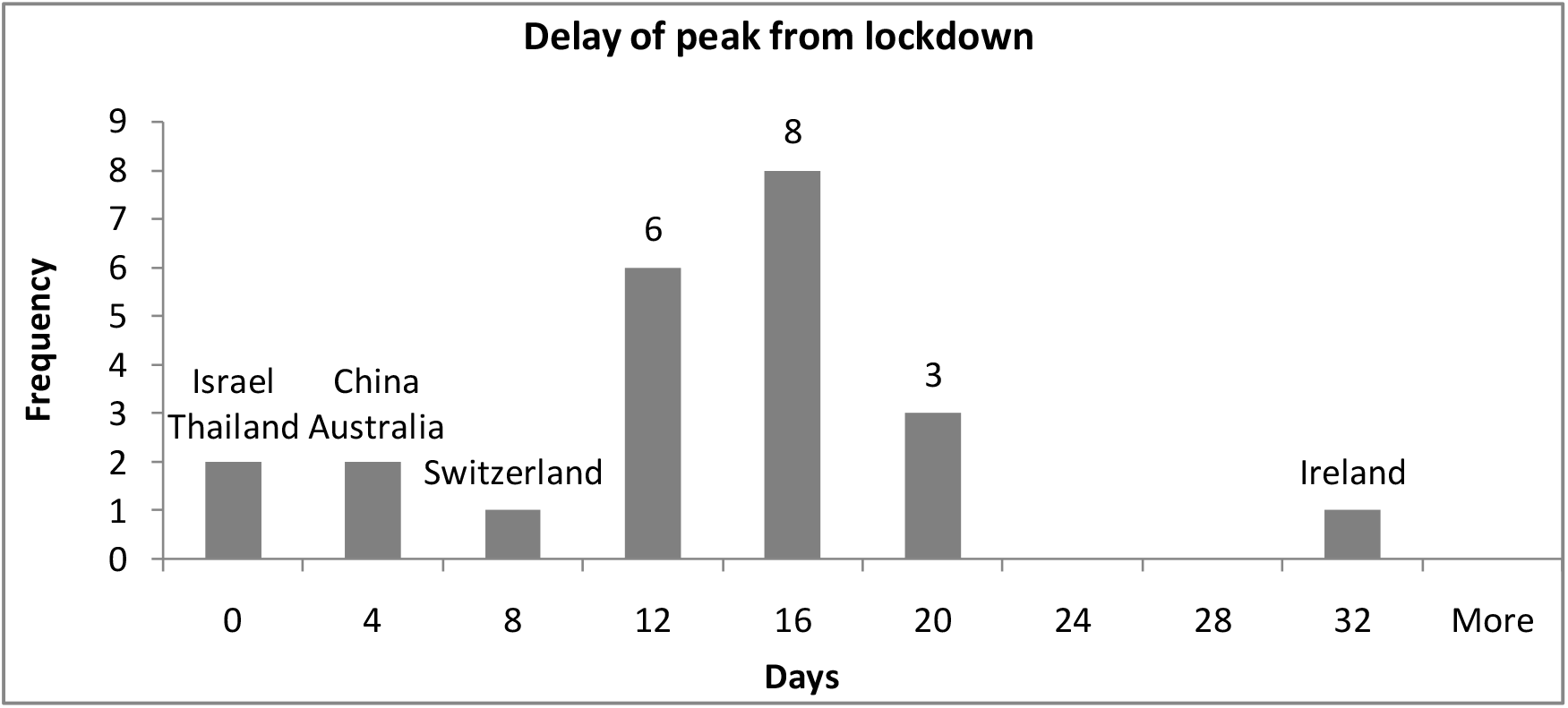
A histogram of the delay between lockdown start date and subsequent peak of daily cases for the countries studied. Seventeen countries reached a peak within 8 to 19 days from lockdown start.

The logistic parameters of the early S-curves carry greater uncertainties than those of the S-curves of the overall fits. From the simulation study of Debecker and Modis we know that typical uncertainties for these early curves are ±20% on *M*1, ±5% on *α*1, and ±1 day on *t*1_0_, all with 90% confidence level. These uncertainties are small enough to permit us to make meaningful comparisons between the early S-curve and the overall S-curve. The difference between these two curves reflects the interventions by the government in question.

### 3.1 On the Rapidity of Government Actions

Governments began taking action, typically a lockdown, when the number of infections in their country reached a certain threshold, say a level *p*, which we can evaluate using Equation (1) as follows:

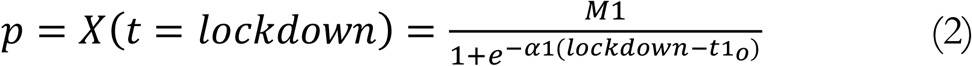

It is more appropriate to use this value of *p* instead of the actual number of infections on that day because daily fluctuations (of statistical origin and not only) dilute the natural aspect of this threshold.

Wikipedia lists the dates of lockdown starts for twenty-four of our twenty-five countries (for the Rest of USA—i.e. without New York—lockdown was approximated as an average weighted by the populations of nine major states.)[15] It is of interest to compare the threshold among different countries. In Figure 4 we see that 70% of the countries we studied reacted with a lockdown before the confirmed cases in their country reached 10,000. The remaining 7 countries—with the exception of Israel—are the countries where the virus spread profusely; more on this in Section 3.4 below.

**Figure 4.**
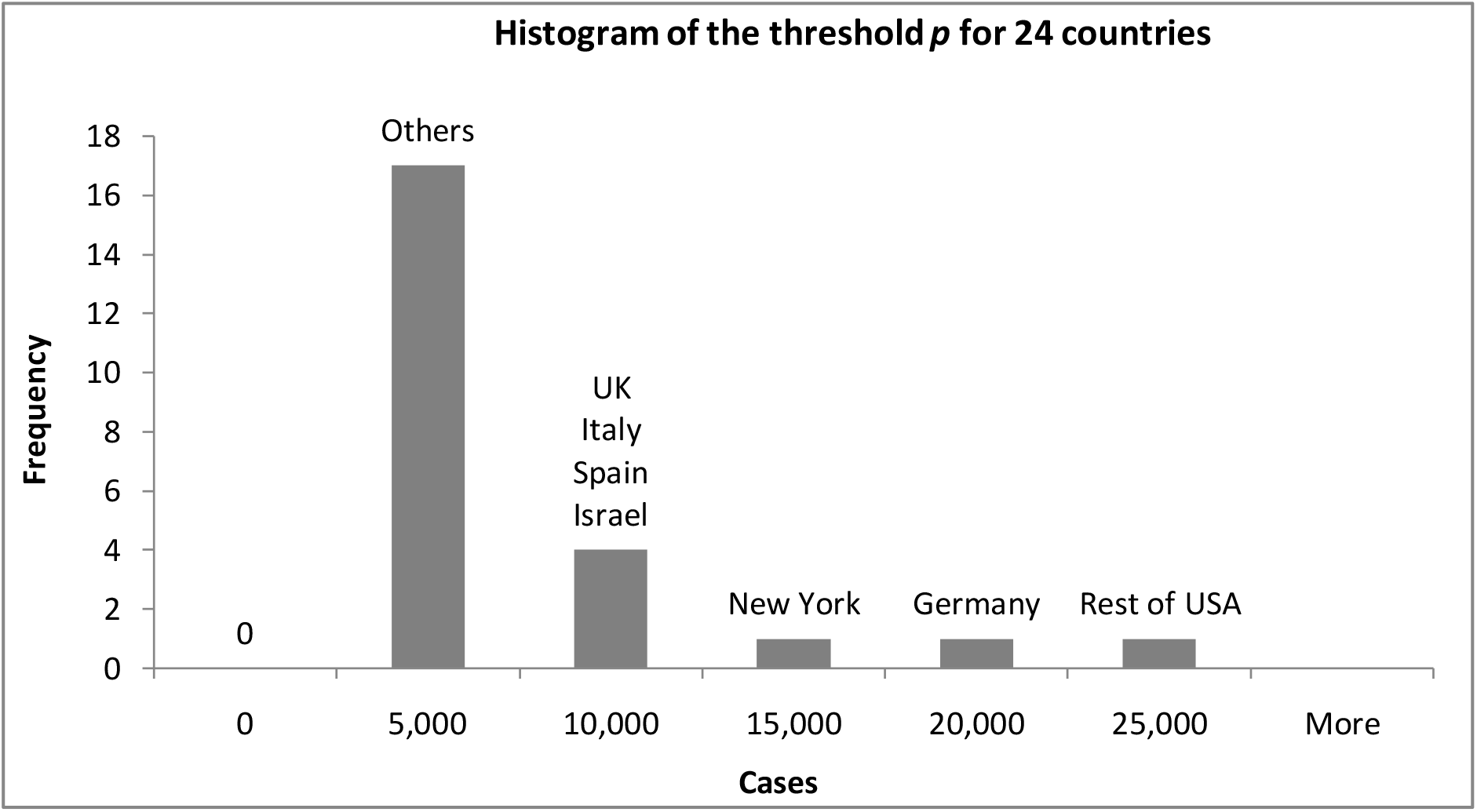
A histogram of the threshold *p* —the number of confirmed cases at lockdown start—for the countries studied.

A further observation was that a low threshold *p* is associated with a high diffusion rate *α*1, see scatter plot in Figure 5. This could be understood as follows: a more rapid early spreading of the virus alarmed governments more and triggered a response (e.g. lockdown) at a lower number of infections.

**Figure 5.**
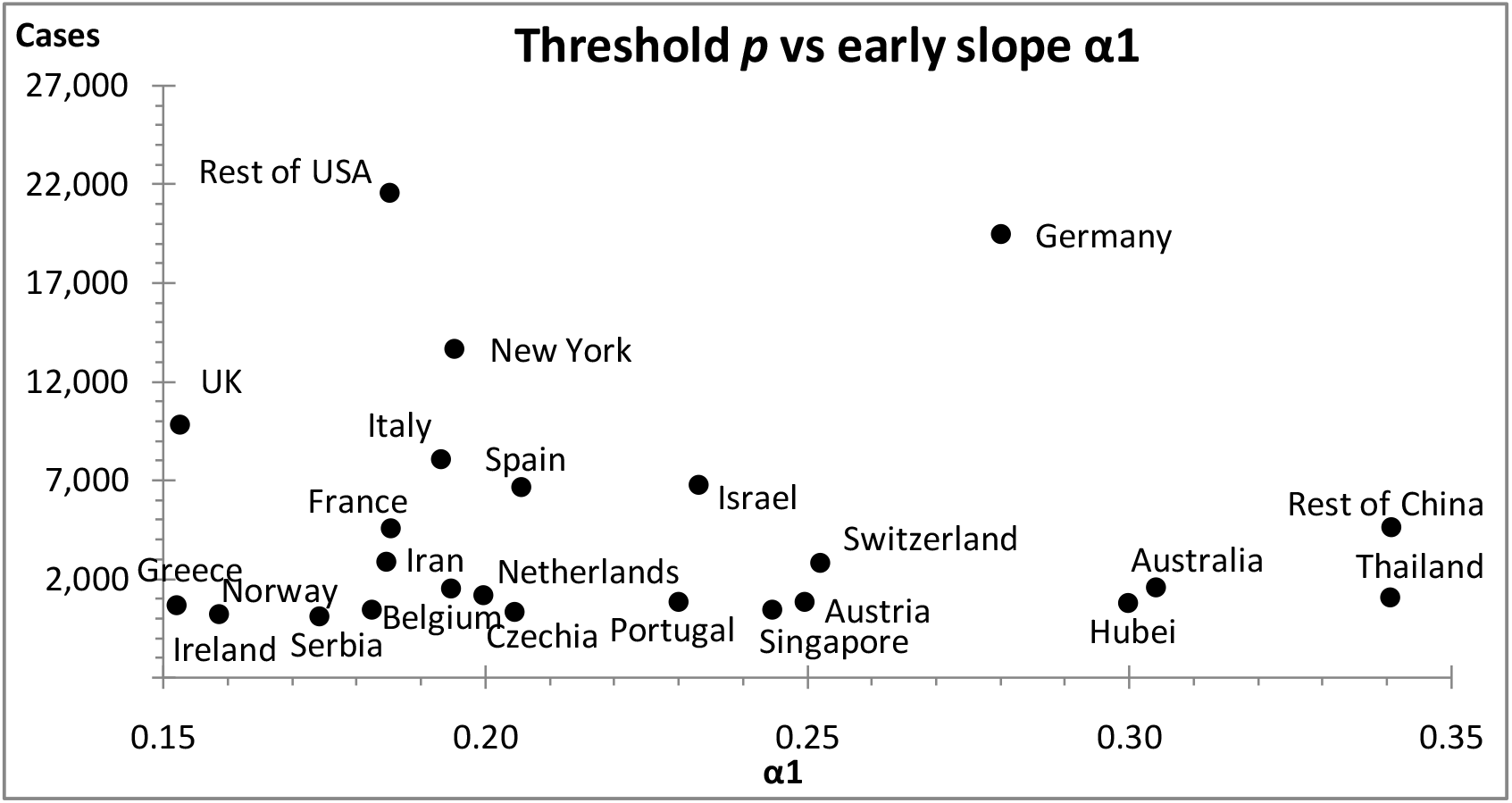
A scatter plot of the slope *α*1 of the early S-curves versus the threshold *p*; high diffusion rates are preferentially associated with low thresholds.

### 3.2 On the Efficiency of Flattening the Curve

The final curve is flatter than the early curve (i.e. *α*1 > *α*) and the difference (*α*1 – *α*) becomes a measure of the amount of flattening (remember, the smaller the value of, the flatter the curve will be.) Figure 6 shows that in general a greater threshold resulted in a flatter curve. Early reaction—i.e. low values—will flatten less and consequently will distort less the overall logistic S-curve. Thailand seems to be an outlier here, but Thailand was also an outlier in Figure 3 with its lockdown following instead of preceding the peak of daily cases!

**Figure 6.**
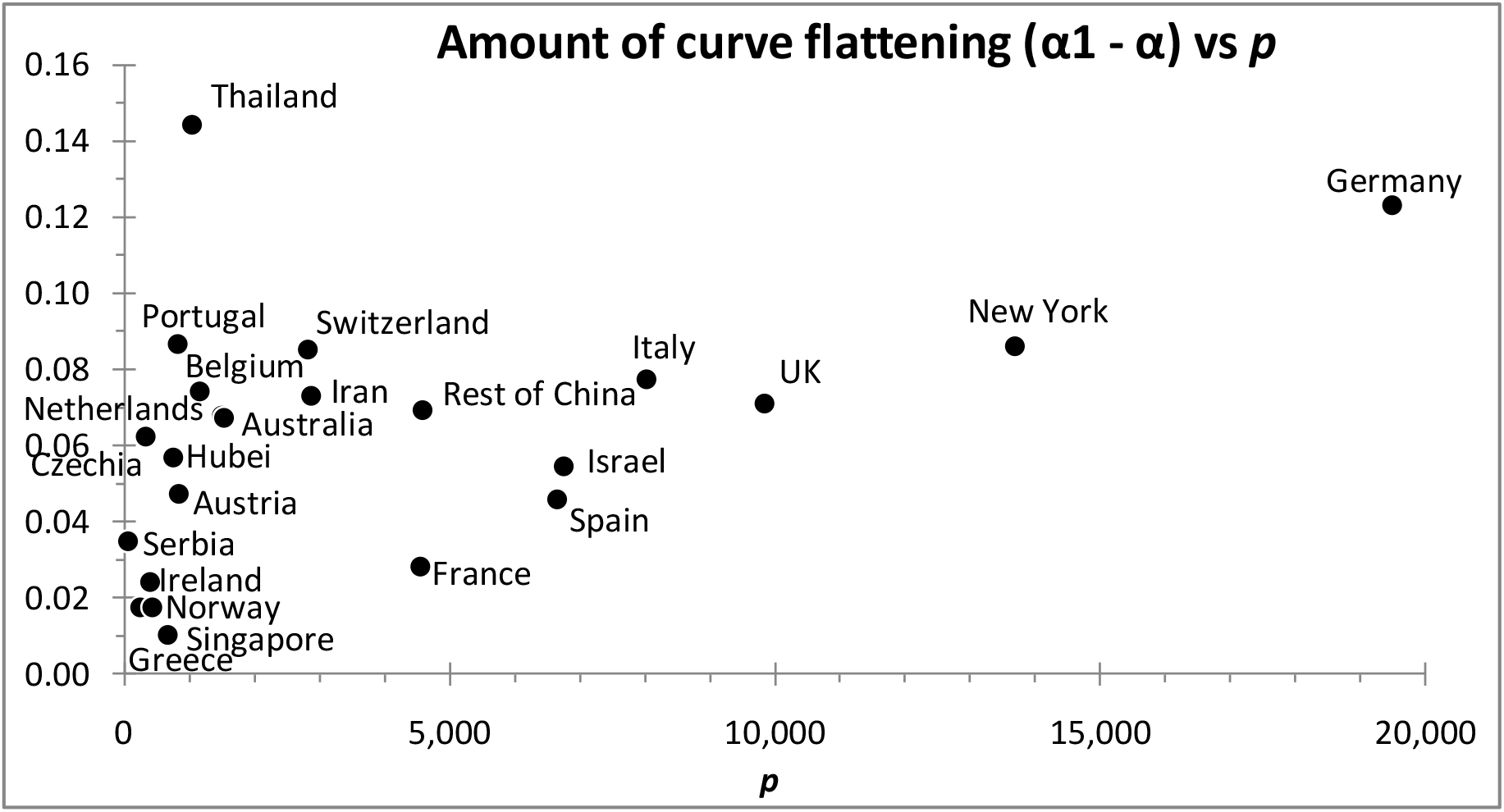
A scatter plot of the amount of curve flattening versus the lockdown threshold *p* ; the higher the threshold, the greater the flattening (and the distortion) of the S-curve.

### 3.3 The Prolongation of the Epidemic

A consequence of flattening the curve is that the midpoint of the S-curve will be delayed. Following government actions the difference between the midpoints of the early and the final S-curves (*t*_0_- *t*1_0_) measures this delay. If we define the percentage reduction of the curve’s slope as*Δα*=(*α*1- *α*)/ *α*1, then the scatter plot in Figure 7 shows that in general the greater the flattening of the curve (i.e. the greater the reduction in) the longer the prolongation of the epidemic, which also results into a more asymmetric final distribution, but more importantly into a greater number of infections, see Section 3.4 below.

**Figure 7.**
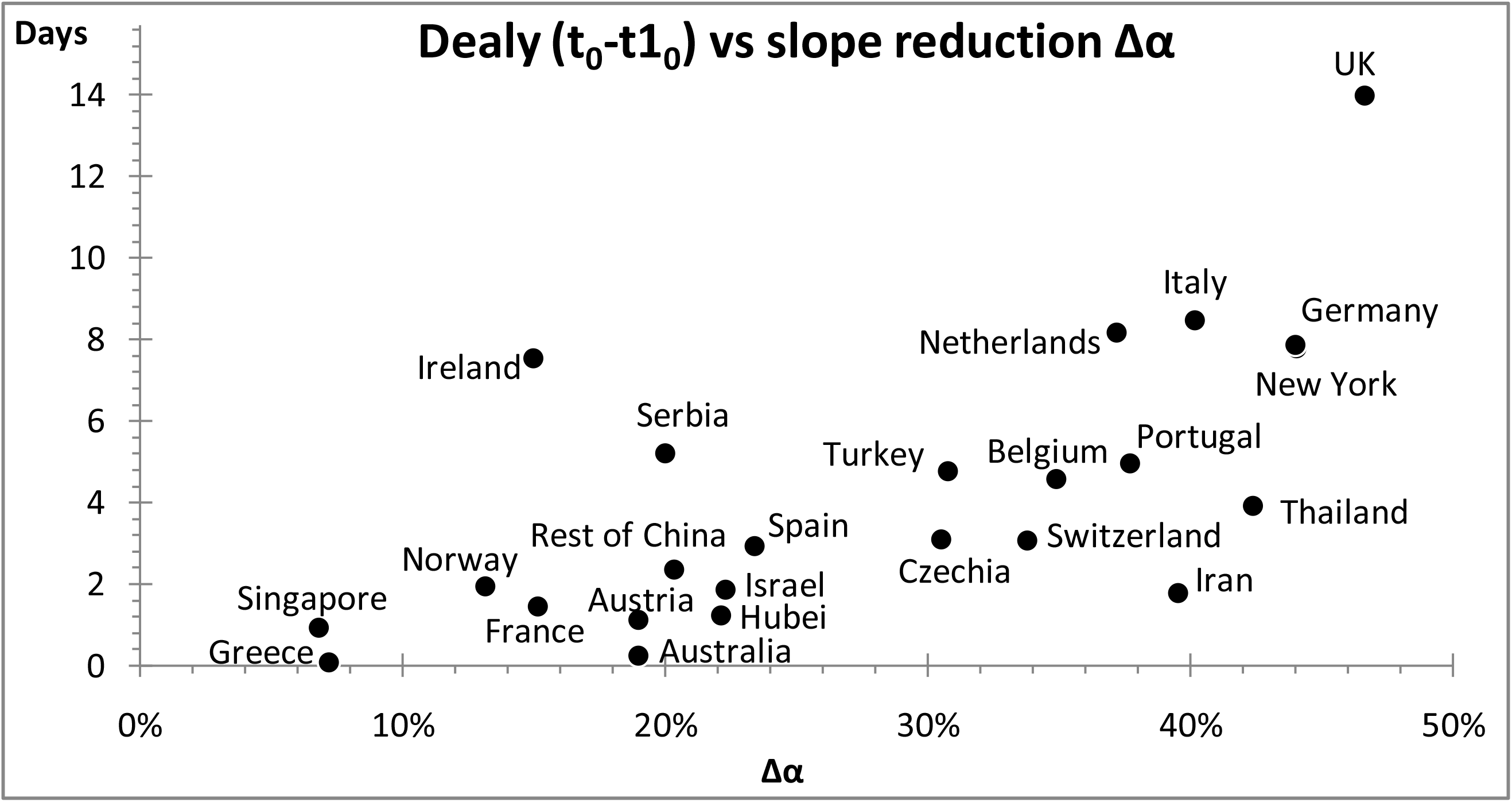
A scatter plot of the delay (*t*_0_- *t*1_0_) versus the percentage reduction in the rate of diffusion ; the greater the flattening, the longer the prolongation.

### 3.4 Increasing the Total Number of Infections

The prolongation of the epidemic (*t*_0_- *t*1_0_) will result in an excess of infections. The ceiling *M* of the overall S-curve is greater than the ceiling *M*1 of the early S-curve and the difference (*M* - *M*1) measures how much the total number of infections is increased by the flattening process. Figure 8 shows that the more the epidemic is prolonged the greater this excess will be.

**Figure 8.**
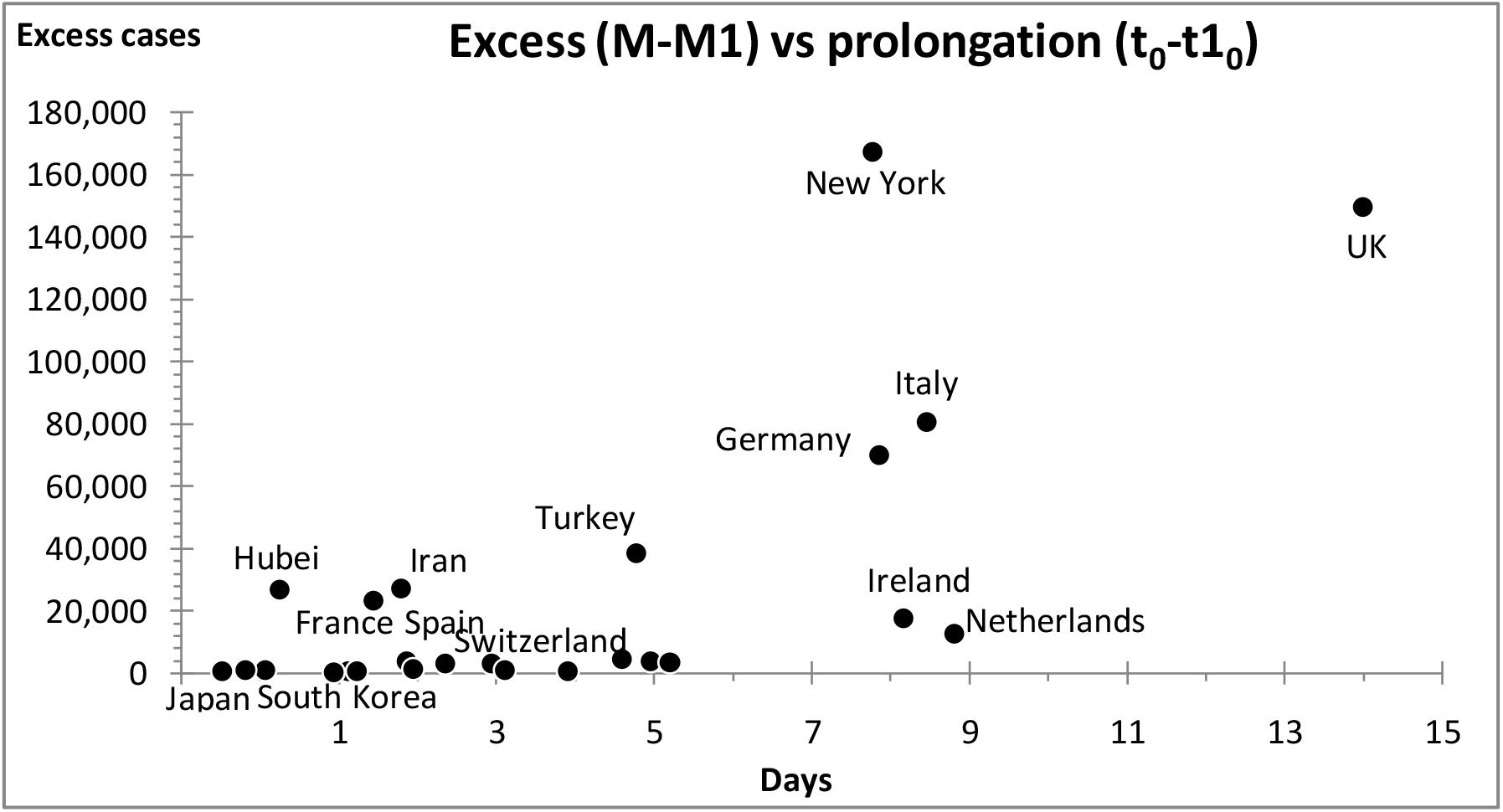
A scatter plot of the excess cases (*M* - *M*1) versus the delay (*t*_0_- *t*1_0_); the greater the prolongation, the greater the excess

It is not surprising that the excess in the number of infections also correlates to the amount of slope reduction *Δα*. In the scatter plot of Figure 9 we see that the greater the amount of flattening the greater the excess will be. This message corroborates the correlation initially observed in Figure 2.

**Figure 9.**
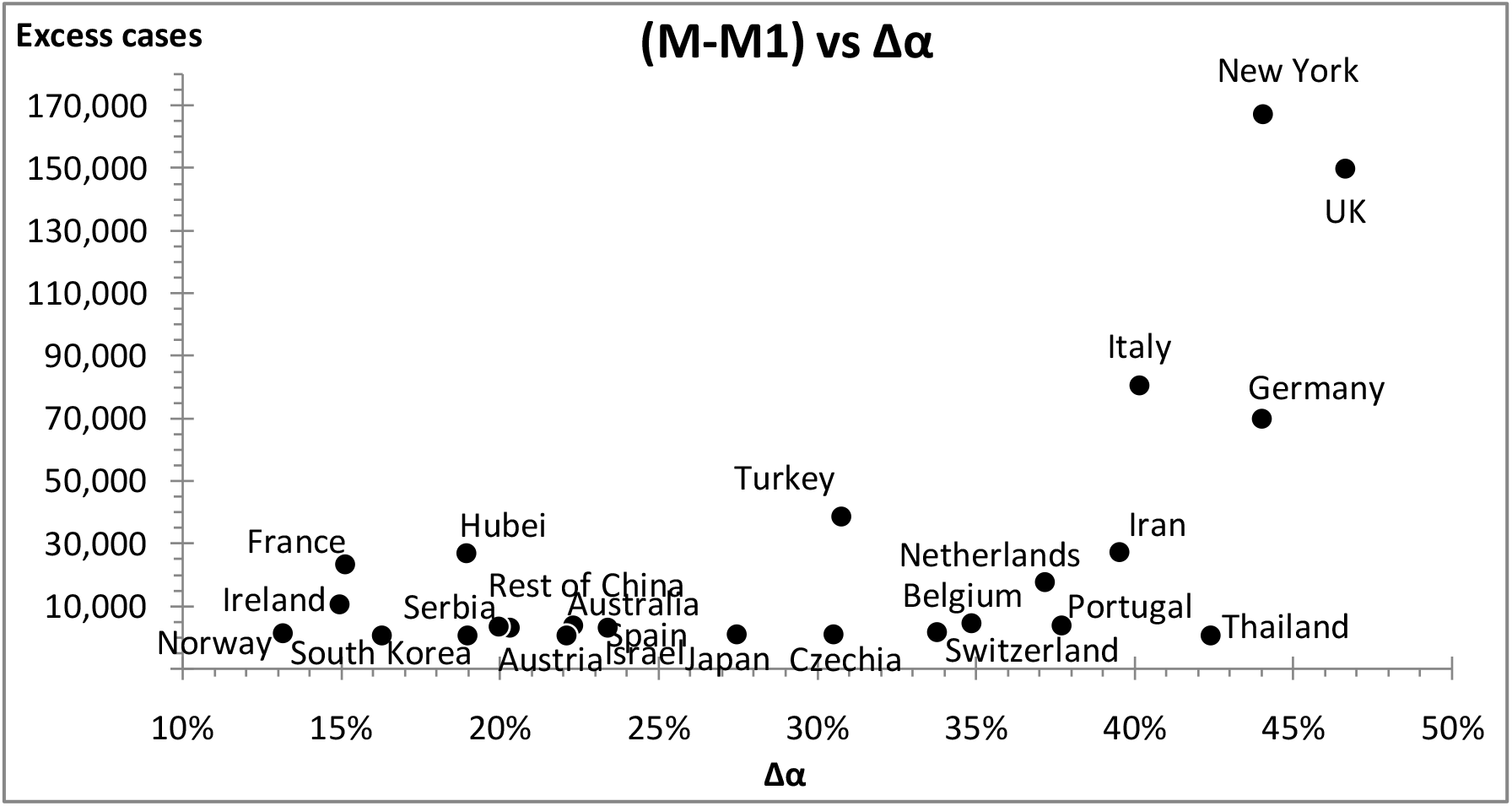
A scatter plot of the excess cases (*M* - *M*1) following flattening of the curve versus the amount of flattening ; the greater the flattening *Δα*;the greater the excess.

But the excess (*M* - *M*1) also increases with the threshold *p*. Figure 10 shows that the higher this threshold the greater the excess.

**Figure 10.**
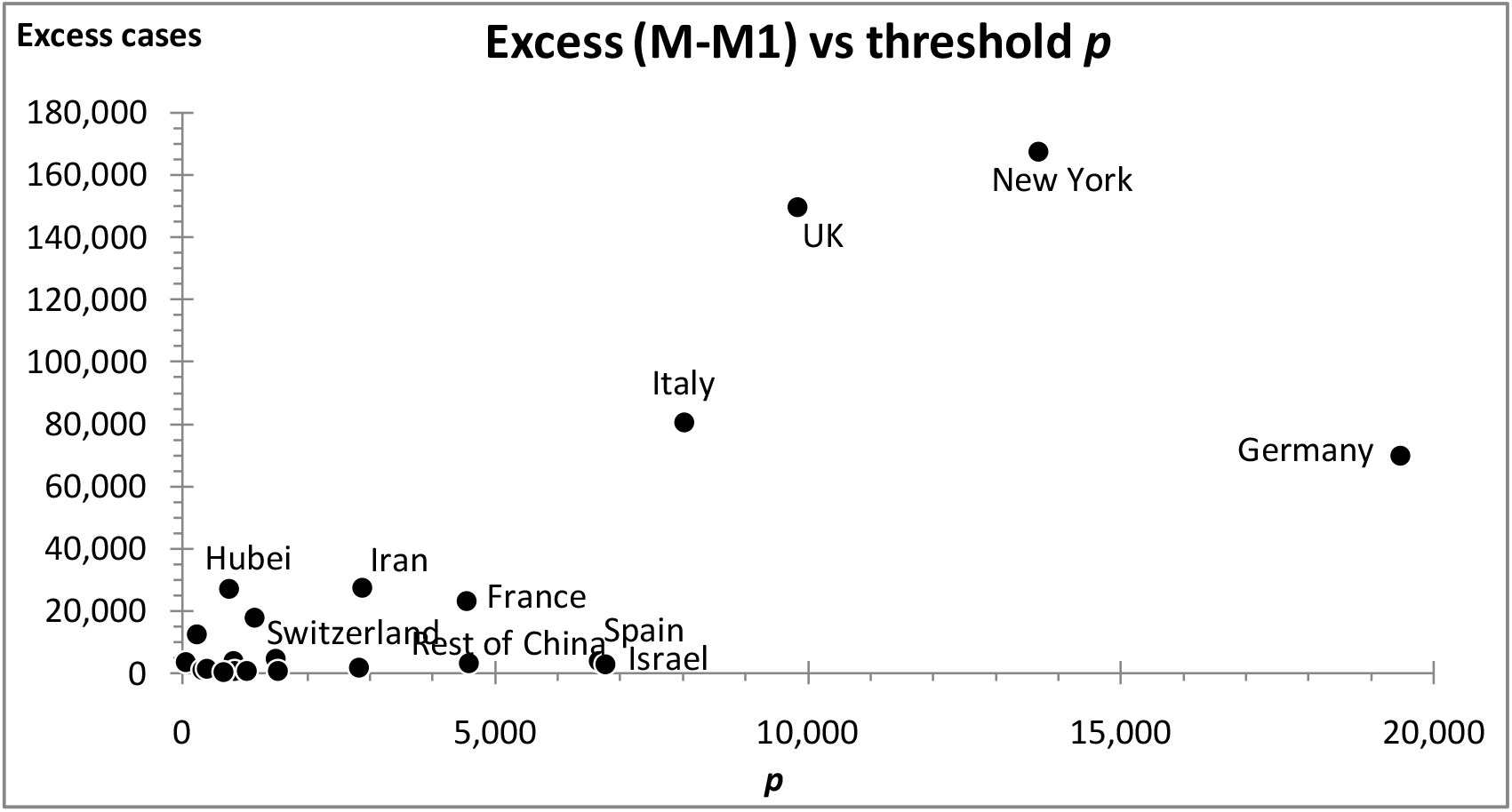
A scatter plot of the excess cases (*M* - *M*1) versus the lockdown threshold ; the higher the threshold, the greater the excess.

## 4. Discussion

The COVID-19 pandemic obliged governments around the world to impose lockdowns in order to slow down the rate of the virus diffusion. They took this action when the number of confirmed cases in their country reached a certain threshold *p*. In so doing they flattened the curve, which began taking effect about two weeks later.

But flattening the curve also prolonged the diffusion process. The midpoint of the curve was delayed; the greater the flattening, the longer this prolongation, which eventually resulted in a greater number of infections.

Countries that responded late (i.e. at a higher threshold *p*) ended up with flatter curves and greater number of infections (e.g. New York, UK, Germany.) In contrast, countries where the virus initially diffused at a higher rate, responded early, i.e. at lower threshold *p* (e.g. Hubei Province, Thailand, Australia,) and ended up with a small increase on the number of infections. Similarly, the steeper the early S-curve (i.e. the larger *α*1) the shorter the prolongation of the epidemic and the more symmetric the final distribution (e.g. South Korea, Rest of China, Australia.) The flattening of the curve produced an asymmetric life- cycle pattern with increased final number *M* of infections in the country (e.g. New York, UK, Italy.) This increase, which is bigger the higher the threshold s*p* and the longer the peak is delayed, corroborates the negative correlation between *M* and *α* that we established in our simulation study decades ago.

Do all these observations imply that flattening the curve was not necessarily the best thing to do? An increased number of infections entails an increased number of deaths. On the other hand, a flatter curve saves lives by avoiding exceeding the capacity of intensive care units in hospitals. Is it possible that more lives were lost by flattening the curve? There is a rather complicated optimization problem here with calculations difficult to carry out quantitatively. There is a moral issue, however, that cannot be overlooked: no civilized society will ever opt out from providing critical care to any fraction of its population.

There are lessons to be learned from the countries that managed well, such as Hubei Province, Rest of China, Austria, and South Korea. These countries acted decisively and swiftly, which resulted in steeply-rising symmetric overall S-curves for the virus diffusion there. The first three instituted lockdowns early, well before seeing 5,000 cases. South Korea did not lockdown but introduced early what was considered one of the largest and best- organized epidemic control programs, with various measures to screen the mass population for the virus, and isolate any infected people as well as trace and quarantine those who contacted them. These countries did not flatten the curve; they simply squeezed it, by limiting the number of potential infections.

In conclusion, the best strategy would be to act early and decisively thus minimizing the distortion of the epidemic’s natural-growth pattern—the S-curve. In so doing there will be minimal flattening and minimal prolongation of the curve; consequently minimal excess of infections. The rate of infections that grew sharply will also decline sharply resulting in a short, symmetric overall natural-growth curve.

## Data Availability

All data used can be found either in the references and/or in text of the manuscript itself.

https://systems.jhu.edu/research/public-health/ncov/

https://www.worldometers.info/

https://data.europa.eu/euodp/en/home

https://en.wikipedia.org/wiki/National_responses_to_the_COVID-19_pandemic

http://creativecommons.org/licenses/by/4.0/

## Appendix Logistic Fit Parameters and Graphs

**Table 1.**
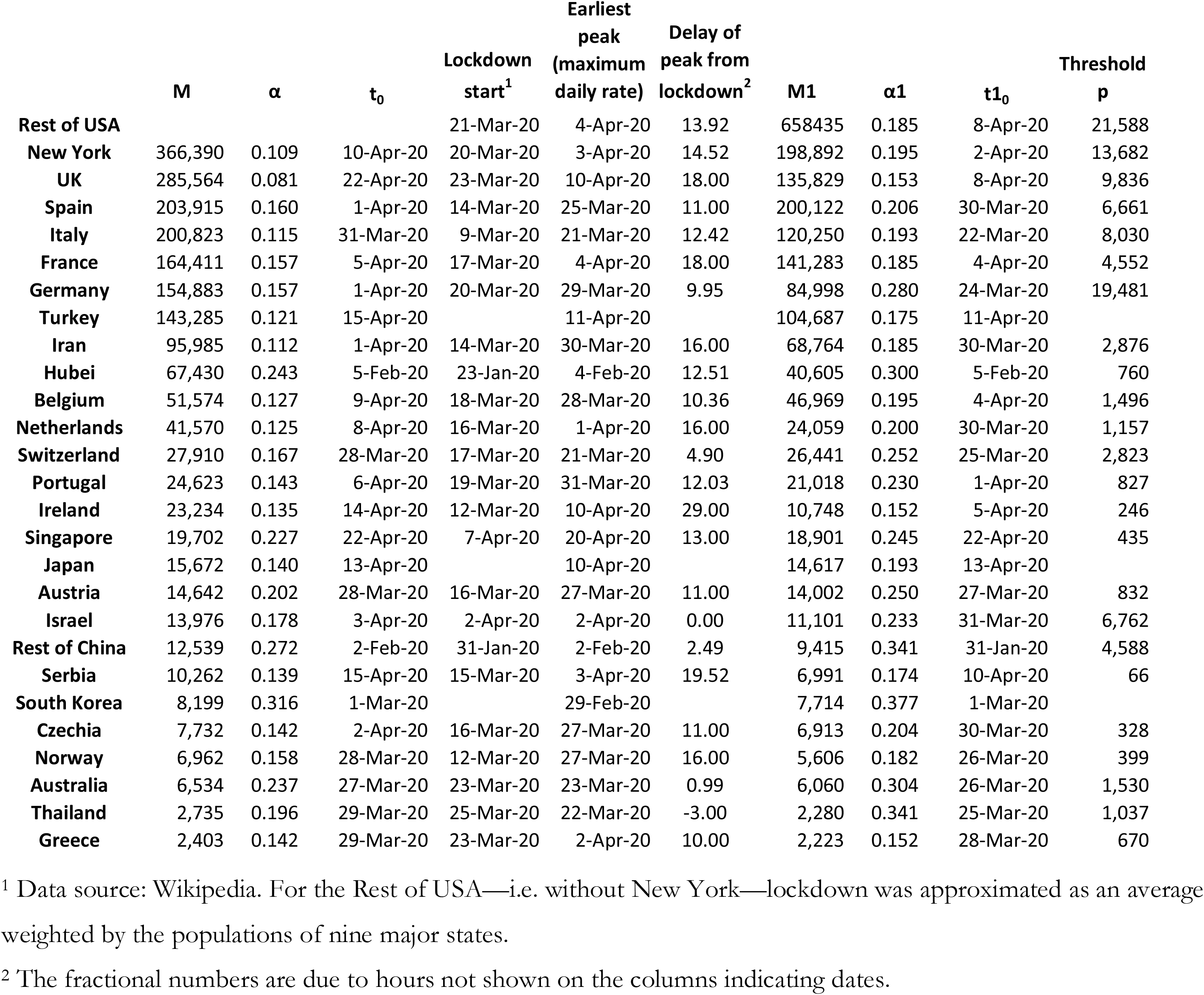
The fit parameters of the overall and the early S-curves and other data.

| | M | $\alpha$ | $t_0$ | Lockdown<br>start<sup>1</sup> | Earliest<br>peak<br>(maximum<br>daily rate) | Delay of<br>peak from<br>lockdown<sup>2</sup> | M1 | $\alpha_1$ | $t_{1_0}$ | Threshold<br>p |
| --- | --- | --- | --- | --- | --- | --- | --- | --- | --- | --- |
| **Rest of USA** |  |  |  | 21-Mar-20 | 4-Apr-20 | 13.92 | 658435 | 0.185 | 8-Apr-20 | 21,588 |
| **New York** | 366,390 | 0.109 | 10-Apr-20 | 20-Mar-20 | 3-Apr-20 | 14.52 | 198,892 | 0.195 | 2-Apr-20 | 13,682 |
| **UK** | 285,564 | 0.081 | 22-Apr-20 | 23-Mar-20 | 10-Apr-20 | 18.00 | 135,829 | 0.153 | 8-Apr-20 | 9,836 |
| **Spain** | 203,915 | 0.160 | 1-Apr-20 | 14-Mar-20 | 25-Mar-20 | 11.00 | 200,122 | 0.206 | 30-Mar-20 | 6,661 |
| **Italy** | 200,823 | 0.115 | 31-Mar-20 | 9-Mar-20 | 21-Mar-20 | 12.42 | 120,250 | 0.193 | 22-Mar-20 | 8,030 |
| **France** | 164,411 | 0.157 | 5-Apr-20 | 17-Mar-20 | 4-Apr-20 | 18.00 | 141,283 | 0.185 | 4-Apr-20 | 4,552 |
| **Germany** | 154,883 | 0.157 | 1-Apr-20 | 20-Mar-20 | 29-Mar-20 | 9.95 | 84,998 | 0.280 | 24-Mar-20 | 19,481 |
| **Turkey** | 143,285 | 0.121 | 15-Apr-20 |  | 11-Apr-20 |  | 104,687 | 0.175 | 11-Apr-20 |  |
| **Iran** | 95,985 | 0.112 | 1-Apr-20 | 14-Mar-20 | 30-Mar-20 | 16.00 | 68,764 | 0.185 | 30-Mar-20 | 2,876 |
| **Hubei** | 67,430 | 0.243 | 5-Feb-20 | 23-Jan-20 | 4-Feb-20 | 12.51 | 40,605 | 0.300 | 5-Feb-20 | 760 |
| **Belgium** | 51,574 | 0.127 | 9-Apr-20 | 18-Mar-20 | 28-Mar-20 | 10.36 | 46,969 | 0.195 | 4-Apr-20 | 1,496 |
| **Netherlands** | 41,570 | 0.125 | 8-Apr-20 | 16-Mar-20 | 1-Apr-20 | 16.00 | 24,059 | 0.200 | 30-Mar-20 | 1,157 |
| **Switzerland** | 27,910 | 0.167 | 28-Mar-20 | 17-Mar-20 | 21-Mar-20 | 4.90 | 26,441 | 0.252 | 25-Mar-20 | 2,823 |
| **Portugal** | 24,623 | 0.143 | 6-Apr-20 | 19-Mar-20 | 31-Mar-20 | 12.03 | 21,018 | 0.230 | 1-Apr-20 | 827 |
| **Ireland** | 23,234 | 0.135 | 14-Apr-20 | 12-Mar-20 | 10-Apr-20 | 29.00 | 10,748 | 0.152 | 5-Apr-20 | 246 |
| **Singapore** | 19,702 | 0.227 | 22-Apr-20 | 7-Apr-20 | 20-Apr-20 | 13.00 | 18,901 | 0.245 | 22-Apr-20 | 435 |
| **Japan** | 15,672 | 0.140 | 13-Apr-20 |  | 10-Apr-20 |  | 14,617 | 0.193 | 13-Apr-20 |  |
| **Austria** | 14,642 | 0.202 | 28-Mar-20 | 16-Mar-20 | 27-Mar-20 | 11.00 | 14,002 | 0.250 | 27-Mar-20 | 832 |
| **Israel** | 13,976 | 0.178 | 3-Apr-20 | 2-Apr-20 | 2-Apr-20 | 0.00 | 11,101 | 0.233 | 31-Mar-20 | 6,762 |
| **Rest of China** | 12,539 | 0.272 | 2-Feb-20 | 31-Jan-20 | 2-Feb-20 | 2.49 | 9,415 | 0.341 | 31-Jan-20 | 4,588 |
| **Serbia** | 10,262 | 0.139 | 15-Apr-20 | 15-Mar-20 | 3-Apr-20 | 19.52 | 6,991 | 0.174 | 10-Apr-20 | 66 |
| **South Korea** | 8,199 | 0.316 | 1-Mar-20 |  | 29-Feb-20 |  | 7,714 | 0.377 | 1-Mar-20 |  |
| **Czechia** | 7,732 | 0.142 | 2-Apr-20 | 16-Mar-20 | 27-Mar-20 | 11.00 | 6,913 | 0.204 | 30-Mar-20 | 328 |
| **Norway** | 6,962 | 0.158 | 28-Mar-20 | 12-Mar-20 | 27-Mar-20 | 16.00 | 5,606 | 0.182 | 26-Mar-20 | 399 |
| **Australia** | 6,534 | 0.237 | 27-Mar-20 | 23-Mar-20 | 23-Mar-20 | 0.99 | 6,060 | 0.304 | 26-Mar-20 | 1,530 |
| **Thailand** | 2,735 | 0.196 | 29-Mar-20 | 25-Mar-20 | 22-Mar-20 | -3.00 | 2,280 | 0.341 | 25-Mar-20 | 1,037 |
| **Greece** | 2,403 | 0.142 | 29-Mar-20 | 23-Mar-20 | 2-Apr-20 | 10.00 | 2,223 | 0.152 | 28-Mar-20 | 670 |

<sup>1</sup> Data source: Wikipedia. For the Rest of USA—i.e. without New York—lockdown was approximated as an average weighted by the populations of nine major states.
<sup>2</sup> The fractional numbers are due to hours not shown on the columns indicating dates.

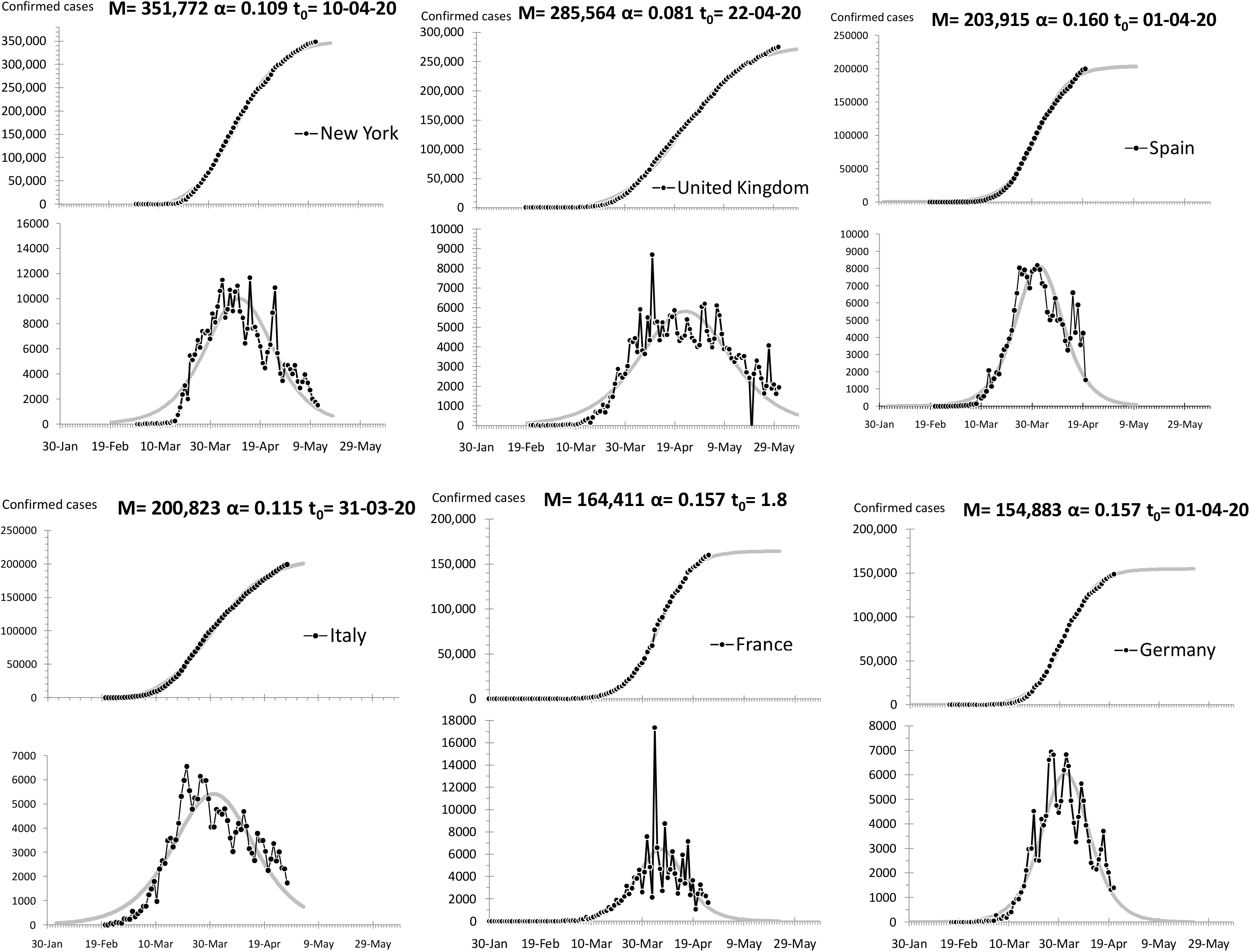

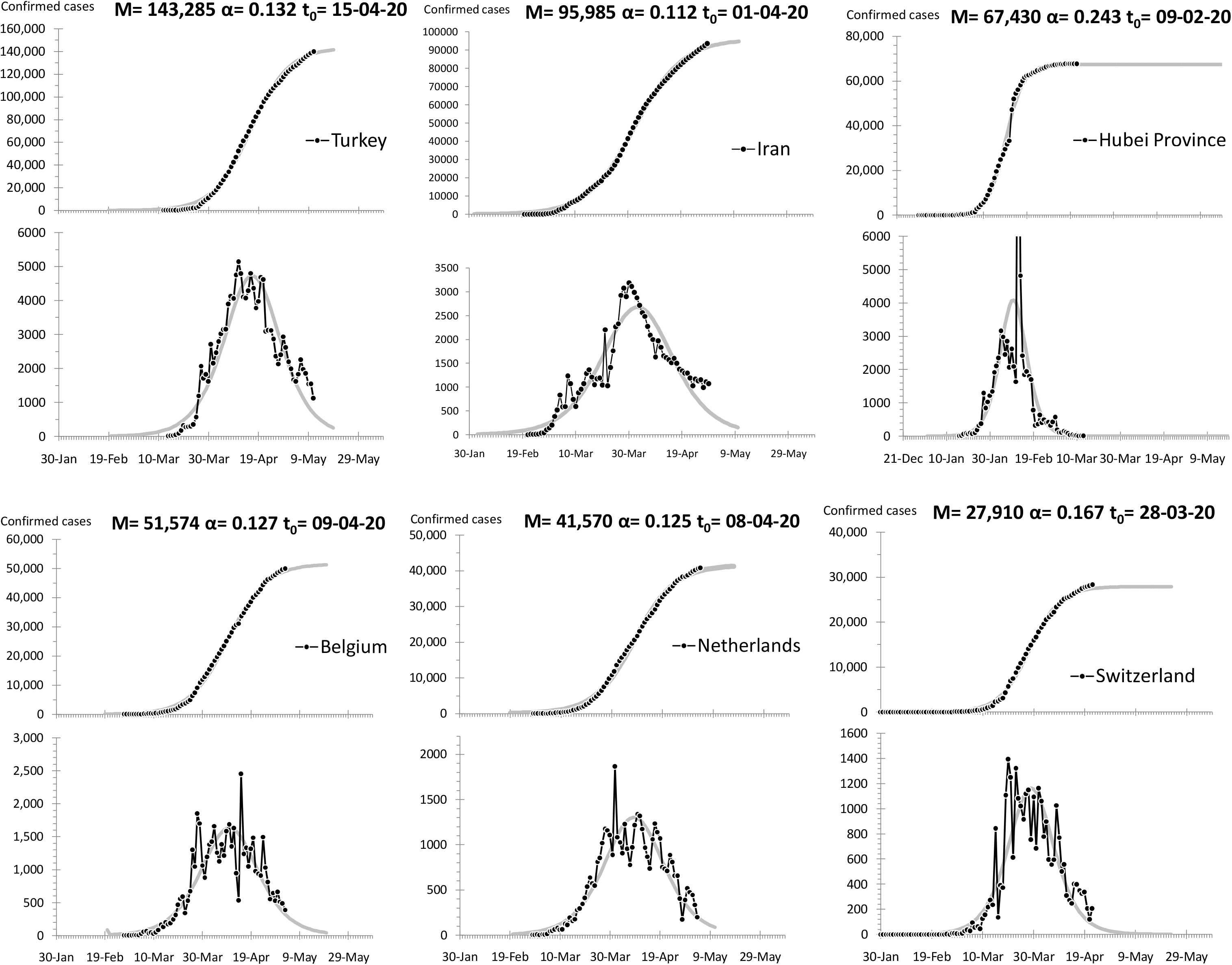

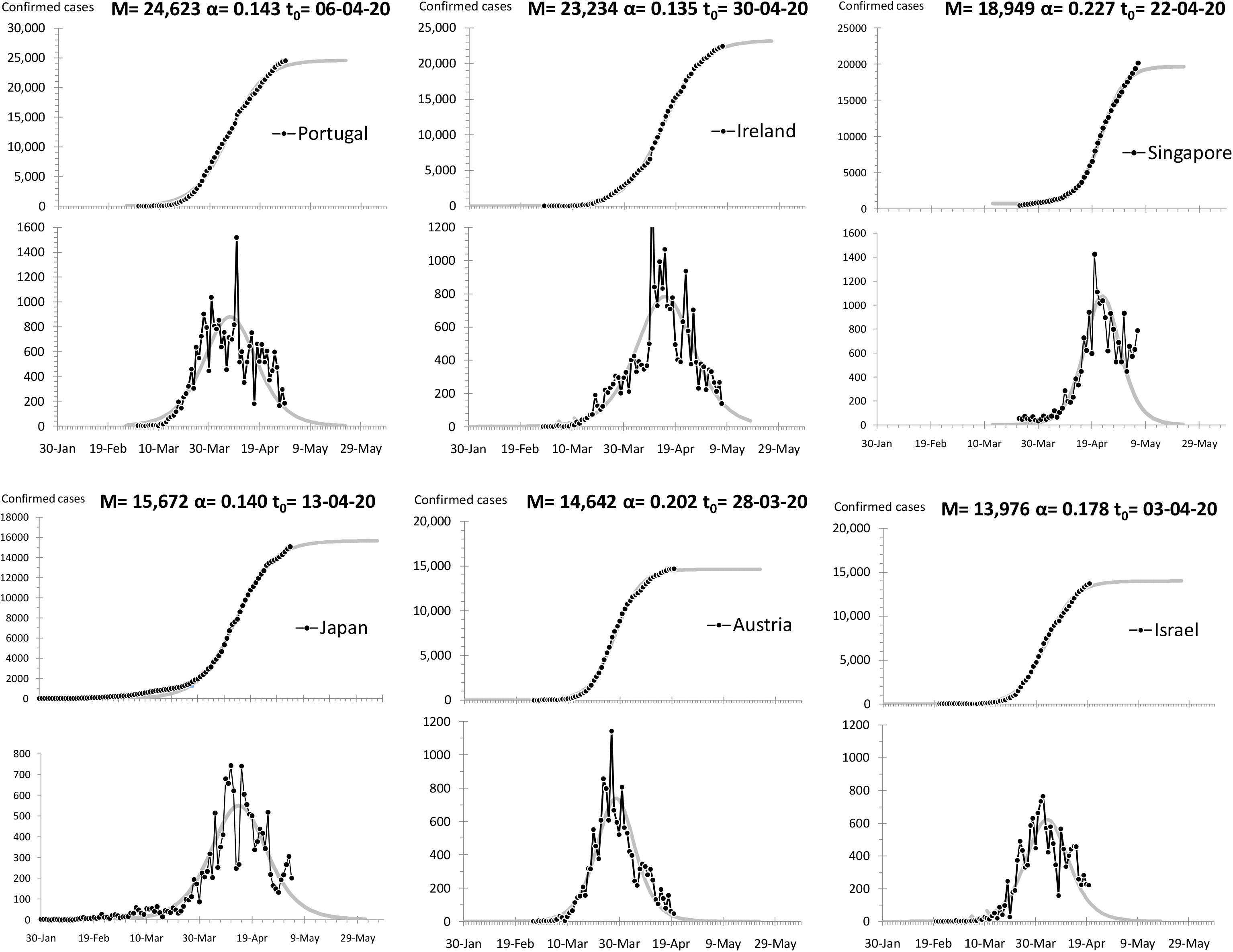

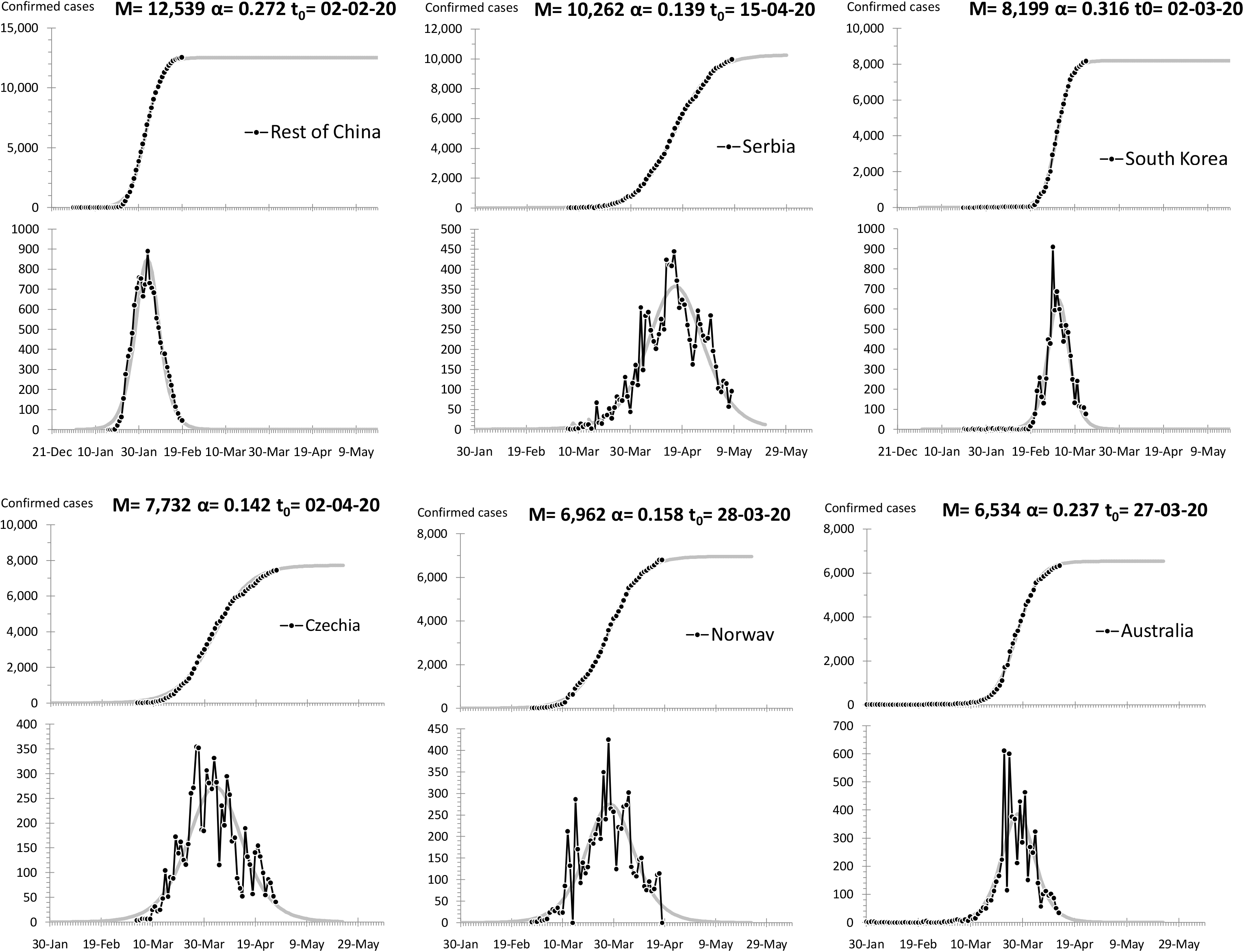

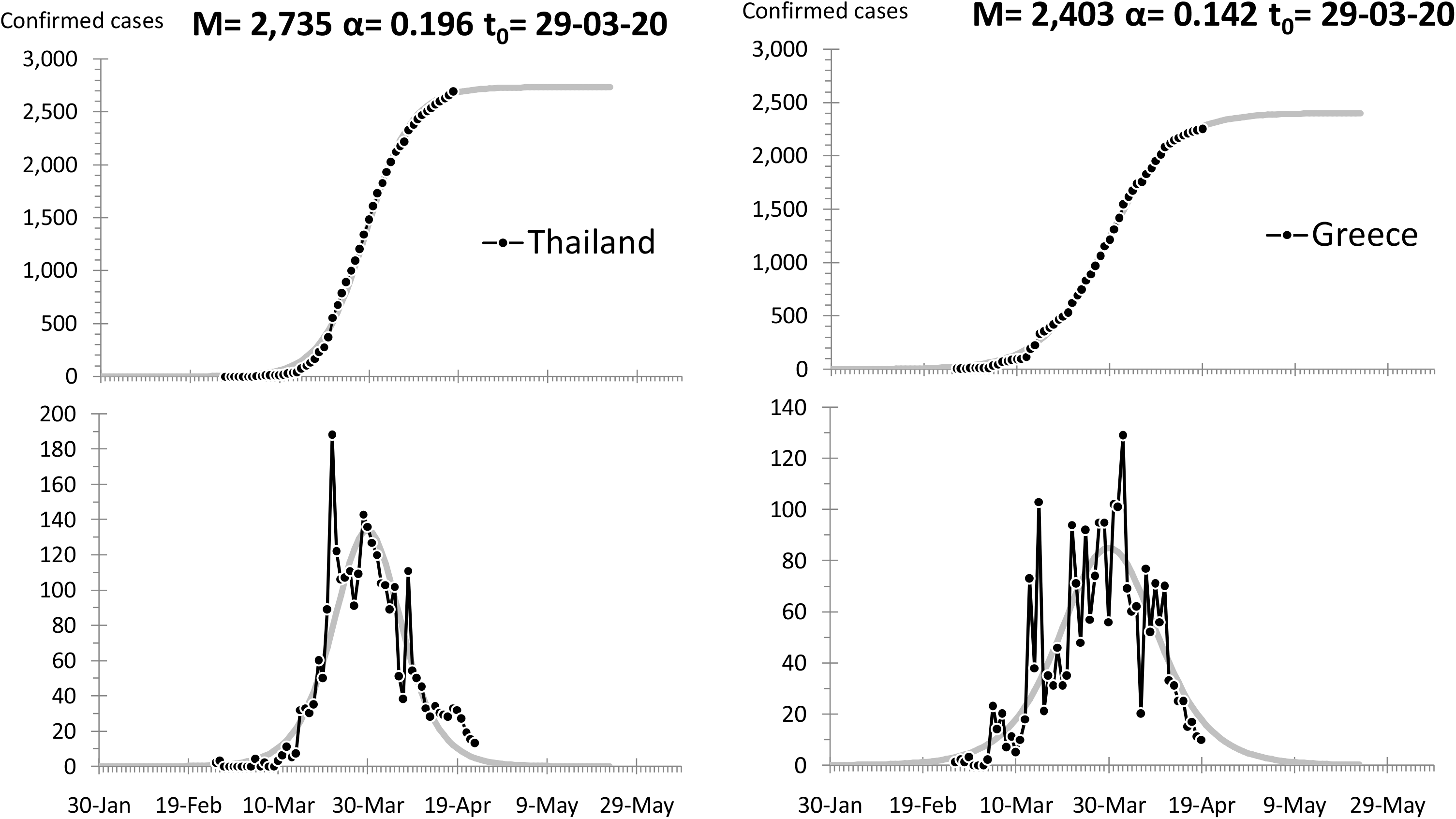

*One of us, Theodore Modis, thanks Professor Athanasios G. Konstantopoulos, Chairman of the Board, Center for Research and Technology Hellas, for fruitful discussions*.

## Biographies

Alain Debecker is a mathematician and data scientist working at IMAD [Institut de Maintien à Domicile], Geneva. During the previous ten years he was associate professor in Management Science at the University of Lyon. He has also made Business modeling and data analysis at Digital Equip. Corp (DEC), the State of Geneva, the United Nations, and others.

Theodore Modis is a physicist, strategist, futurist, and international consultant. He is author/co-author to about one hundred articles in scientific and business journals and ten books. He has on occasion taught at Columbia University, the University of Geneva, at business schools INSEAD and IMD, and at the leadership school DUXX, in Monterrey, Mexico. He is the founder of Growth Dynamics, an organization specializing in strategic forecasting and management consulting: http://www.growth-dynamics.com

